# Worries about COVID-19 infection and psychological distress at work and while commuting

**DOI:** 10.1101/2021.02.16.21250657

**Authors:** Masamichi Uehara, Tomohiro Ishimaru, Hajime Ando, Seiichiro Tateishi, Hisashi Eguchi, Mayumi Tsuji, Koji Mori, Shinya Matsuda, Yoshihisa Fujino, for the CORoNaWork Project

## Abstract

**Objective:** This study examined the relationship between worry about COVID-19 infection in the workplace and while commuting to work and psychological distress in Japan.

**Methods:** An internet monitor study was conducted. Out of a total of 33,302 participants, 26,841 people were included. The subjects were asked single-item questions about whether they were worried about COVID-19 infection in general, at work and while commuting to work. Kessler 6 (K6) was used to assess psychological distress.

**Results:** The OR was significantly higher in association with worry about infection in the workplace at 1.71 (95%CI 1.53–1.92) and worry about infection while commuting at 1.49 (95%CI 1.32–1.67).

**Conclusions:** This study suggests the need for psychological intervention to reduce worry about infection in response to public mental health challenges associated with the COVID-19 pandemic.

## Introduction

Mental health problems associated with the COVID-19 pandemic are an emerging public health issue^1^. Outbreaks increase generalized fear, community anxiety, and panic symptoms^2,3^. Previous studies have reported a higher prevalence of psychological symptoms such as depression and anxiety among individuals who experienced lockdown during the COVID-19 pandemic ^1,4–7^

Containment measures against pandemics like COVID-19 have a strong impact on individuals’ daily lives and psychological well-being^8^. “A new normal” of avoiding the “three Cs” (crowded places, close-contact settings, and confined and enclosed spaces) has been recommended as an effective countermeasure against infection^8^. However, such measures also distance people from each other, which can lead to decreased communication and socializing. In addition, COVID-19 has also brought about economic recession and job insecurity. Together, these effects are thought to have contributed to an increase in loneliness, worry, fear, anxiety, and stress among individuals.

In situations like the current pandemic, worry of infection is a common feeling among the majority of the population, given that various aspects of daily life pose potential risk of infection. An increase in infections can trigger psychological symptoms such as generalized fear and widespread community anxiety^2,3^. Psychological symptoms such as persistent worrying, feeling overwhelmed by emotions, restlessness and irritability emerge within the population in the form of anxiety, panic attacks, depression and suicide^9,10^. Worry of infection is an early sign of psychological distress.

For workers, the workplace and commuting^11–14^ are considered particularly risky opportunities for infection. For this reason, most workplaces are taking various infection control measures, including limiting the number of visitors, restricting business trips, encouraging telecommuting, ensuring sufficient office ventilation, and installing partitions in customer-facing spaces.^15,16^ However, some workplaces are not taking sufficient recommended measures for business reasons or due to cost.^17,18^ Naturally, infection control efforts in the workplace influence workers’ worry of infection. When infection control does not meet expectations, workers’ anxiety and depression are high.^19^ In contrast, intensive workplace measures against COVID-19 are associated with lower psychological distress among employees.^15,19^ In addition, anxiety and depression are higher among workers who do not telecommute and those with interpersonal tasks.^19^

Therefore, we hypothesized that worry of infection in the workplace and while commuting affect the psychological distress of workers. In this study, we examined the relationship between workers’ worry about infection and psychological distress in the midst of a rapid outbreak of COVID-19 in Japan.

## Method

### Study design and subjects

This cross-sectional, internet monitor study was conducted on December 22–26, 2020, when Japan experienced its third wave of COVID-19 infection. Details of the protocol of this survey are reported elsewhere^20^. Briefly, data were collected from workers who had employment contracts at the time of the survey and were selected based on prefecture, job type, and sex.

Out of a total of 33,302 people who participated in the survey, 27,036 people were included in the study after removing those found to have provided fraudulent responses. Participants were excluded if they had exceptionally short response times (≤6 minutes), exceptionally low body weight (<30 kg), exceptionally short height (<140 cm), provided inconsistent responses to comparable questions (e.g., inconsistent responses to questions on marital status and area of residence), and gave incorrect responses to a question included to identify fraudulent responses (select the third largest number from among the following five numbers). After further excluding 195 individuals who indicated in the survey that they had already been infected with COVID-19, a total of 26,841 individuals (13,713 males and 13,128 females) were included in the current analysis. This study was approved by the ethics committee of the University of Occupational and Environmental Health, Japan (reference No. R2-079). Informed consent was obtained in the form of the website.

### Assessment of worry about COVID-19 infection

Three single-item questions were used to determine whether or not the subjects were worried about infection. One question inquired about general worry of COVID-19 infection, while the other two were situation dependent, inquiring about worry of infection at work and while commuting to work. The questions were, “Are you worried about being infected with COVID-19?” “Are you worried about being infected while working at your workplace?” and “Are you worried about being infected while commuting to work?” The participants responded “yes” or “no” to the questions.

### Assessment of psychological distress

Kessler 6 (K6) was used to assess psychological distress^21^. The validity of the Japanese version of the K6 has been confirmed^22^. In the present study, a K6 score of 5 or higher was used as the cutoff for mild psychological distress, and a score of 13 or higher as the cutoff for severe psychological distress.

### Other covariates

We selected potential confounding factors which are possibly related to anxiety and psychological distress. The following survey items were considered confounding factors: age, sex, marital status (married, unmarried, bereaved/divorced), occupation (mainly desk work, jobs mainly involving interpersonal communication, and mainly labor), number of employees, educational background, equivalent income (household income divided by the square root of household size), smoking status, alcohol consumption, frequency of telecommuting, and use of public transportation when commuting. The questionnaire also asked the following questions: “Have you been a close contact of someone infected with COVID-19?” and “Do you know of anyone close to you (friends or family) who has been infected with COVID-19?”

The questionnaire also asked the participants to rate their company’s infection control measures using the question “Do you think your company has taken adequate infection control measures for its employees?” Participants responded on a four-point scale: “yes,” “somewhat,” “not really,” “no.”

In addition, the cumulative incidence rate of COVID-19 infection one month prior to conduct of the survey in the prefectures of residence was used as a community-level variable. Information was collected from the websites of public institutions.

### Statistical analysis

The odds ratios (ORs) of psychological distress associated with worry about infection were estimated using a multilevel logistic model nested in the prefectures of residence. Psychological distress was defined as a K6 score of 5 or higher and 13 or higher.

The multivariate model was adjusted for sex, age, education, equivalent household income, occupation, number of business establishments, smoking status, alcohol consumption, frequency of telecommuting, use of public transportation when commuting, perceived assessment of workplace infection control efforts, presence of infection among acquaintances, and experience of being a close contact. The incidence rate of COVID-19 by prefecture was also used as a prefecture-level variable.

We further estimated the multivariate ORs of psychological distress associated with use of public transportation, with adjustment for all factors except worry about infection during commuting, because adjusting for worry about infection would be an over-adjustment.

A *p* value less than .05 was considered statistically significant. All analyses were conducted using Stata (Stata Statistical Software: Release 16; StataCorp LLC, TX, USA).

## Results

Table 1 shows the characteristics of the subjects according to their worries about infection. A total of 75% of the participants were worried about infection. While 50% of participants were worried about being infected in the workplace, 32% were worried about being infected while commuting to work. Women were more worried about infection than men. There were no substantial differences in income, educational background, or lifestyle by worry about COVID-19 infection in general, while at work, or while commuting. Participants who indicated that they worried about infection in the workplace and while commuting shared similar characteristics.

**Table 1.**
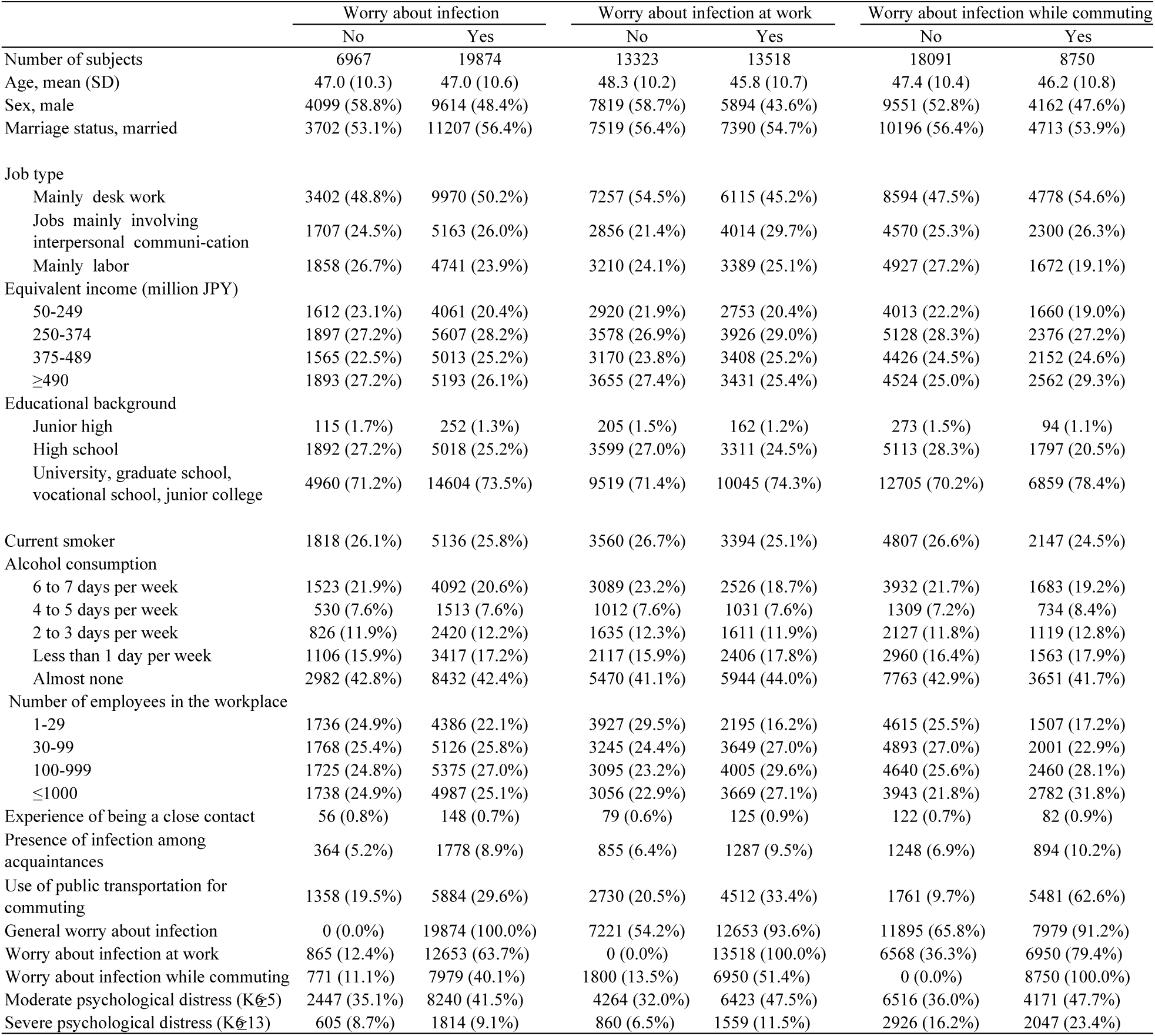
The characteristics of the subjects according to their worries about infection.

Table 2 shows the ORs of psychological distress. In the age-adjusted model for moderate psychological distress defined by a K6 score of 5 or higher, general worry about COVID-19 infection (OR=1.29, 95%CI 1.22–1.37), worry about infection at work (OR=1.79, 95%CI 1.70–1.88), and worry about infection while commuting (OR=1.64, 95%CI 1.55–1.73) all showed significant associations with psychological distress. Multivariate analysis showed that the OR of moderate psychological distress associated with general worry about COVID-19 infection was 0.97 (95% CI 0.91–1.04). The ORs were significantly higher in association with worry about infection in the workplace at 1.57 (95%CI 1.57–1.48) and worry about infection while commuting at 1.49 (95%CI 1.38–1.60).

**Table 2.**
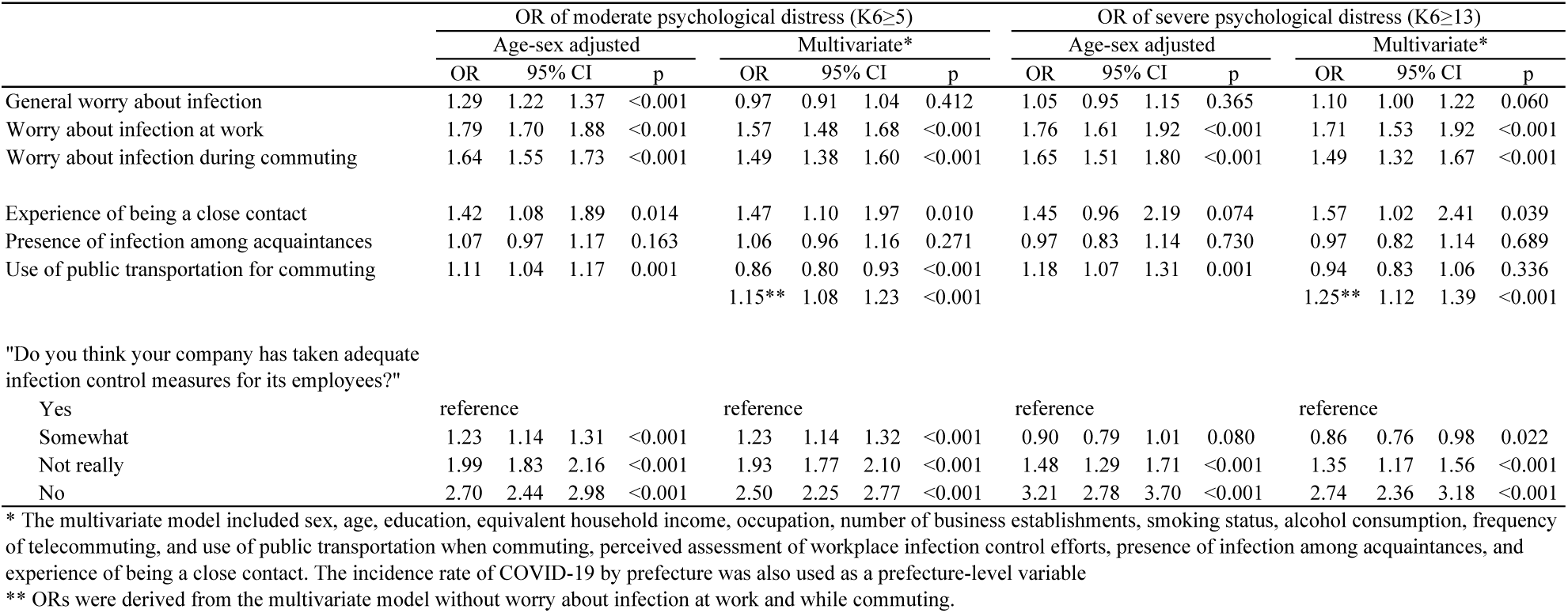
Associations of worry about infection and psychological distress

The sex- and age-adjusted OR of severe psychological distress, defined by a K6 score of 13 or higher, associated with general worry about COVID-19 infection was 1.05 (95%CI 0.95–1.15). ORs were significantly higher in association with worry about infection in the workplace (OR=1.76, 95%CI 1.61–1.92) and worry about infection while commuting (OR=1.65, 95%CI 1.51–1.80). Multivariate analysis of severe psychological distress showed similar results.

We also examined situations we predicted could enhance worry. Being in close contact with an infected person increased the ORs of severe psychological distress; however, having acquaintances who had been infected was not significantly associated with severe psychological distress. The OR of severe psychological distress associated with using public transportation for commuting to work was 1.25 (95%CI 1.25–1.39).

## Discussion

This study showed that while general worry about COVID-19 infection was not associated with psychological distress, worry about infection at work and while commuting were associated with moderate and severe psychological distress.

The spread of COVID-19 infection is causing people to worry about infection in a number of everyday situations. In the present study, 75% of participants were worried about infection, suggesting that COVID-19 infection is a major source of public stress. However, in this situation, worrying about infection is not unhealthy, but rather, a natural reaction to current events. Such worry about infection may lead to preventive behaviors^23–25^. In this situation, we found that having general worry about COVID-19 infection did not lead to psychological distress.

However, our study indicates that certain everyday situations, such as working in the workplace and commuting to work, are particularly worrisome in relation to infection. We found that worry about infection at work was associated with moderate and severe psychological distress. There are many opportunities in the workplace for direct contact with others, such as while communicating with colleagues and providing services to customers, and for indirect contact with people through objects. Previous studies have also reported an increase in anxiety and depression in interpersonal work.^19^ Additionally, in places like offices, workers share a relatively small space. Thus, the fact that individuals have limited control over maintaining the recommended three Cs strategy for infection prevention at work can be a major cause of stress. A sense of security and trust that workplaces can be made safe in the event of a pandemic may counteract any negative feelings.^18^

The present study also showed that worry about infection during commuting was associated with moderate and severe psychological distress. This result is consistent with previous studies that found a higher prevalence of anxiety and depressive symptoms among workers who have to commute to work.^18,19^ Commuting to work is considered one way in which risk of infection can increase^26^, because individuals come into contact with an unspecified number of people, are unable to maintain social distancing, and are present in a space that feels relatively enclosed, such as a bus or train. In fact, 47% of participants who were worried about infection while commuting experienced moderate psychological distress, and 23% experienced severe psychological distress. These results suggest that reducing worries about infection in the workplace and during commuting is important for reducing public mental health issues caused by COVID-19^27^.

However, our findings also suggest the difficulty of reducing psychological distress due to worry about infection. We found that worries about infection in the workplace and while commuting to work were associated with psychological distress even after adjusting for assessment of adequate infection control measures in the workplace and use of public transportation. This result is consistent with a previous study that showed that workers’ anxiety about infection differs in different workplaces, and cannot be explained by workplace-related COVID-19 risk management strategies.^18^ These results suggest that workers worry regardless of whether or not they use public transportation to commute to work or have adequate infection prevention measures installed in their workplace, and that these worries are related to psychological distress. In fact, 38% of those who were worried about their commute did not use public transportation. While we found that public transportation use was associated with psychological distress, this association disappeared after adjusting for general worry about COVID-19 infection. This finding supports the idea that worry, rather than facts, is associated with psychological distress. Worry about infection is not only based on rational judgment, but is also an intuitive feeling. Therefore, to reduce excessive worry about infection and the resulting psychological distress, it is necessary to both promote infection prevention measures and adjust individuals’ perception of their worries.

Recent studies have provided useful suggestions for reducing anxiety about COVID-19 infection. Establishing infection control policies and implementing infection control measures in the workplace can be beneficial for the mental health of workers, and a sense of trust and security that infection control measures in the workplace are adequate will alleviate anxiety.^15,18,19^ Use of personal protective equipment when conducting interpersonal tasks and access to testing in case of suspected infectious contact can also ease anxiety.^18,19^ Additionally, public awareness, such as through access to correct information and avoiding excessive exposure to the media, may be useful for reducing anxiety.^28–30^ Cognitive-behavioral therapy is also considered an effective method for such a purpose^31^.

Limitations of this study warrant mention. First, the generalizability of the results is uncertain because this study was conducted through internet monitors. However, we attempted to reduce as much bias in the target population as possible by sampling according to region, job type, and prefecture based on the infection incidence rate. Second, the study asked simple questions about individuals’ worries about infection. Dichotomous yes/no responses can prevent a nuanced understanding of the real sensations experienced by individuals. However, the questions were not intended to diagnose psychiatric symptoms or psychiatric illnesses. Participants who indicated that they were worried may also exhibit psychiatric symptoms, such as anxiety, depression, and panic; however, distinguishing these symptoms was outside the scope of this study. Third, worry about infection in the workplace is thought to be greatly influenced by the type of job and the environment in which one works. Health care workers are a typical example of a population that is at high risk of infection and are more likely to experience psychological distress^10^. In this study, we only adjusted for simple variables such as job type (desk work and physical work) and the number of employees in the workplace. Fourth, because this was a cross-sectional study, the temporal relationship between worry about infection and psychological distress is unclear. Excessive anxiety and depression may be symptoms of psychological distress. Previous studies have shown that people with a history of psychiatric disorders are more anxious and show greater psychological distress during the COVID-19 pandemic^32^. However, because it is easier to confirm feelings of worry than to make a psychiatric diagnosis, we believe that it is useful to identify the presence of worry among workers for determining health status.

In conclusion, while general worry about infection was not associated with psychological distress, worry about infection in specific situations such as in the workplace and while commuting was associated with moderate and severe psychological distress. These worries and the associated psychological and emotional responses may not be mediated by actual infection control measures taken in the workplace or use of public transportation. Thus, despite the continued spread of COVID-19 infection and an increase in anxiety in the population, situational fear, but not generic fear, is correlated with poor mental health. Such knowledge will be useful for designing future policies to manage mental health. The results of this study suggest the need for psychological approaches to reduce worries about infection in response to the mental health challenges associated with the COVID-19 pandemic.

## Data Availability

Data not available due to ethical restrictions.

## Acknowledgements

The current members of the CORoNaWork Project, in alphabetical order, are as follows: Dr. Yoshihisa Fujino (present chairperson of the study group), Dr. Akira Ogami, Dr. Arisa Harada, Dr. Ayako Hino, Dr. Hajime Ando, Dr. Hisashi Eguchi, Dr. Kazunori Ikegami, Dr. Kei Tokutsu, Dr. Keiji Muramatsu, Dr. Koji Mori, Dr. Kosuke Mafune, Dr. Kyoko Kitagawa, Dr. Masako Nagata, Dr. Mayumi Tsuji, Ms. Ning Liu, Dr. Rie Tanaka, Dr. Ryutaro Matsugaki, Dr. Seiichiro Tateishi, Dr. Shinya Matsuda, Dr. Tomohiro Ishimaru, and Dr. Tomohisa Nagata. All members are affiliated with the University of Occupational and Environmental Health, Japan.

